# Associations of inflammation-related proteome with demographic and clinical characteristics of people with HIV in South Africa

**DOI:** 10.1101/2022.12.15.22283410

**Authors:** Junyu Chen, Qin Hui, Chang Liu, Jaysingh Brijkumar, Johnathan A. Edwards, Claudia E. Ordóñez, Mathew R. Dudgeon, Henry Sunpath, Selvan Pillay, Pravi Moodley, Daniel R. Kuritzkes, Mohamed Y. S. Moosa, Tooru Nemoto, Vincent C. Marconi, Yan V. Sun

## Abstract

Elevated levels of inflammation associated with HIV infection are considered one of the primary causes for the excess burden of age-related morbidity and mortality among people with HIV (PWH). Circulating protein levels can be used to investigate biological pathways contributing to persistent inflammation among PWH. In this study, we profiled 73 inflammation-related protein markers and assessed their associations with chronological age, sex and CD4+ cell count among 87 black South African PWH prior to initiating ART. We identified 1, 1 and 14 inflammatory proteins significantly associated with sex, CD4+ T-cell count, and age respectively among PWH. Twelve out of 14 age-associated proteins have been reported to be associated with age in the general population, and 4 have previously shown significant associations with age for PWH. Furthermore, many of the age-associated proteins such as CST5, CCL23, SLAMF1, MMP-1, MCP-1, and CDCP1 have been linked to chronic diseases such as cardiovascular disease and neurocognitive decline in the general population. We also found a synergistic interaction between male sex and older age accounting for excessive expression of CST5. In conclusion, we found that age may lead to the elevation of multiple inflammatory proteins among PWH. We also demonstrated the potential utility of proteomics for evaluating and characterizing the inflammatory status among PWH.

## Introduction

Human immunodeficiency virus (HIV) is a serious public health issue affecting over 38.4 million people worldwide, with an estimated 1.5 million new infections in 2021^1^. With the introduction of effective antiretroviral therapy (ART), HIV-related mortality has been substantially reduced^2^. However, prolonged survival with HIV infection has been associated with an increased risk of age-related comorbidities such as hypertension, cardiovascular diseases, diabetes, and cancers compared to age-matched individuals without HIV infection^3-8^. This health impact is significant for populations with highly prevalent and prolonged HIV epidemics such as the population of South Africa^9^.

The elevated level of inflammation associated with HIV infection is considered one of the primary causes for the excess burden of age-related morbidity and mortality^6^. Even though ART dramatically decreases systemic inflammation, residual inflammation remains a chronic driver of age-related comorbidities for PWH^8,10^. Therefore, assessing the underlying biological pathways contributing to persistent inflammation among PWH with and without ART is of great scientific and public health interest.

Protein biomarkers are measurable molecules that can be used in the clinical setting to diagnose diseases or identify potential therapeutic targets. Many of these measurable proteins are indicative of immune functions, providing new opportunities to study inflammation and biological aging among PWH. Over the past decades, advancement in proteomics profiling techniques over the past decade has enabled the quantification and epidemiologic investigation of thousands of proteins in plasma^11^. Recent studies have pinpointed several inflammatory proteins which were elevated in adults and children with HIV compared to those without HIV independent of the use of ART^6,12^. In addition to HIV infection, many other factors such as age, sex, and CD4+ T-cell counts could lead to differential expression of inflammatory proteins in the general population^13-15^. A recent study investigated how age and sex influenced plasma inflammatory proteins among 192 European PWH, identifying 21 age-associated proteins and 1 sex-associated protein^16^. However, the latter was likely to suffer from insufficient statistical power due to a small number (9%) of female PWH. Furthermore, their results derived from European populations might not apply to other people of other ancestries due to various genetic and environmental factors.

In this study, we profiled 92 inflammation-related protein markers in the plasma to assess their associations with chronological age, sex and CD4+ T-cell count among 87 black South African PWH prior to initiating ART. We focused only on treatment naïve patients because the toxicities induced by ART and the time on treatment can affect aging in PWH^7^, which could complicate the testing and interpretation of phenotype-protein associations in PWH. We identified statistically significant associations of inflammatory proteins which are also linked to chronic diseases such as cardiovascular disease and neurocognitive decline. The results demonstrated the potential values of applying a proteomic approach in studying biological aging and inflammation for PWH.

## Method

### Study population

Our study was based on a sub-cohort from the KwaZulu-Natal (KZN) HIV AIDS Drug Resistance Surveillance Study (ADReSS), which was a prospective study of PWH in care at urban (RK Khan) and rural (Bethesda) HIV clinics in South Africa^17^. Ethical approval from the Biomedical Research Ethics Committee of University of KZN, Emory University, and Mass General Brigham institutional review boards was obtained prior to the start of the study. ART-naïve patients who provided signed informed consent were enrolled just prior to their treatment initiation from 2014 to 2016. Demographic and clinical information including age, sex, race/ethnicity and CD4+ cell count of each ADReSS participant were collected at cohort entry. All participants had blood samples drawn upon enrollment in the study for record of and other circulating biomarkers. Plasma samples were stored in −80 °C and then used to measure the proteomic profile.

### Proteome measures and quality control

Plasma samples obtained from whole blood (EDTA blood tube) were subjected to soluble proteome analysis using the Olink^®^ Targe 96 Inflammation Panel that included 92 proteins linked to immune response or immune-oncology diseases^18^. Breifly, Olink^®^ Targe 96 Inflammation Panel used quantitative PCR (qPCR) to obtain threshold cycle (Ct) values, which is the x-axis value of the point where the reaction curve and the threshold line overlap. Ct values indicate the number of cycles needed for the expression signal to surpass the fluorescent signal threshold line^18^. Normalized Protein Expression (NPX), an arbitrary, relative quantification unit, was derived from Ct values by Olink NPX Signture software:

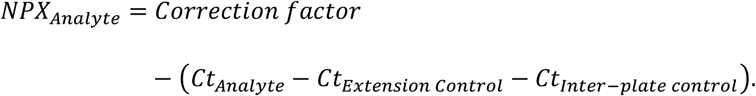

Extension control is used to adjust the Ct values from each sample with respect to extension and amplification. Inter-plate control is a pool of 92 antibodies and is included in triplicate on each plate. The median of the inter-plate control triplicates was used to adjust for potential variation between runs. The correction factor is pre-determined by Olink during the validation of the panels using negative controls. After being adjusted against the correction factor, a difference of 1 NPX approximates a doubling of the protein concentration regardless of protein (log_2_ scale). Limit of detection (LOD) was calculated based on the background estimated from negative controls included on every plate with the following formula:

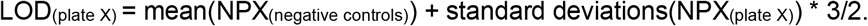

Each sample plate had its own LOD, which is used as the detection threshold (i.e., a protein in a plasma sample is undetectable if its NPX was lower than LOD). Proteins (n=15) that were undetectable in >25% of the samples were excluded from the downstream analysis. Additional 4 proteins (IL4, IL-1-alpha, TSLP, CXCL5) with NPX that did not roughly follow the normal distribution based on Supplement Figure 1 were excluded from downstream analysis. All 87 ADReSS participants with profiled proteomes data passed the internal built-in quality control performed by Olink^®^ NPX Manager software with internal controls.

**Figure 1.**
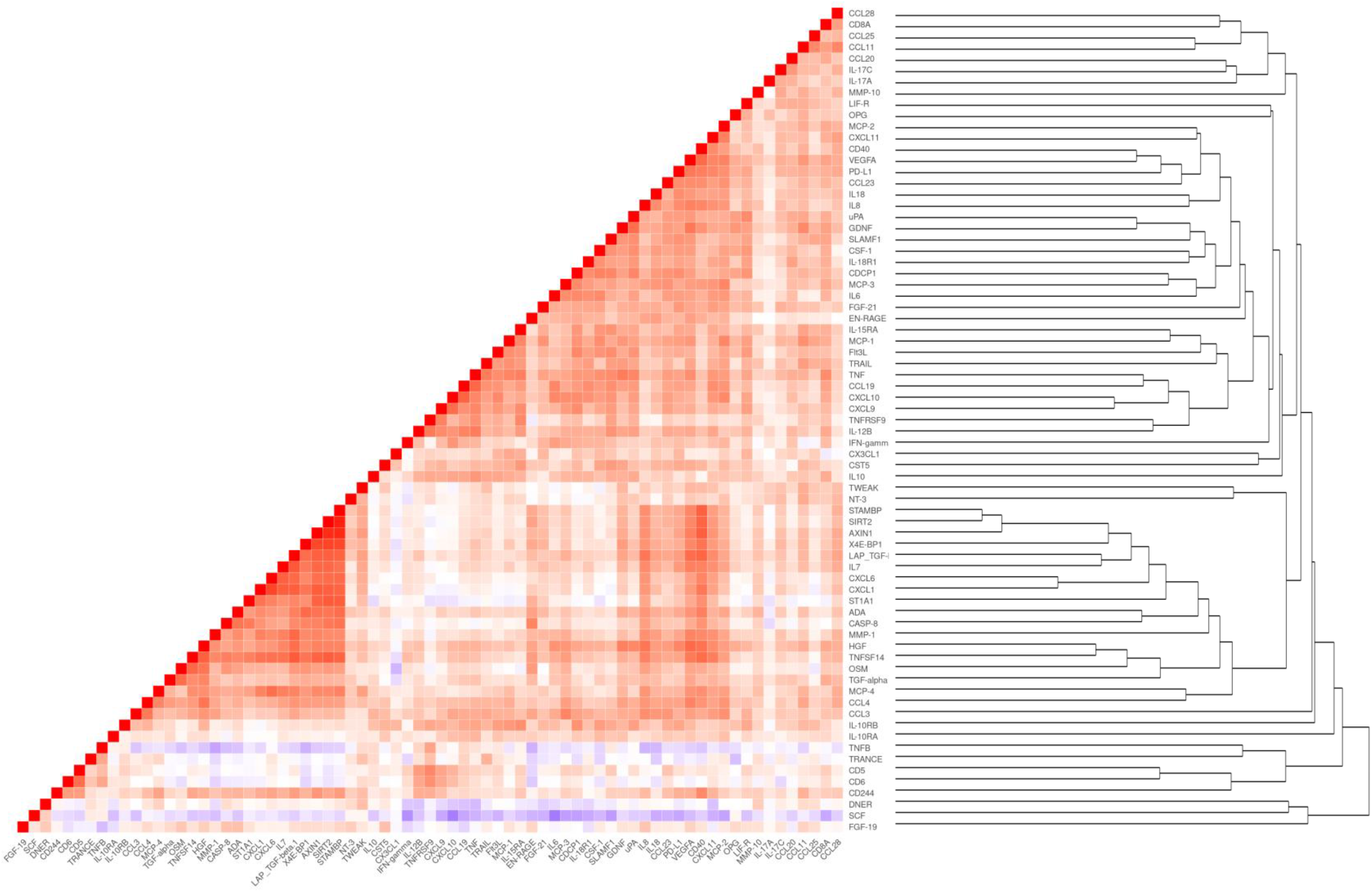
Heatmap showing Spearman correlation between 73 inflammation-related proteins. Dendrogram showing hierarchical clustering of 73 inflammation-related proteins.

### Statistical Analyses

We calculated Spearman correlations to illustrate the relationship between 73 proteins which passed quality control procedures. To examine their similarity, we also performed an agglomerative hierarchical clustering with average-linkage on standardized NPX values of 73 proteins. Linear regression models were used to evaluate the crude associations for all 73 proteins (outcome variables) with 3 traits (sex, age, and CD4+ T-cell count) using proteomes as the outcome. To control for potential confounding, the regression models evaluating the effect of sex and age were adjusted for each other and the study sites. The adjusted regression models evaluating the effect of CD4+ T-cell count were adjusted for sex, age, and study sites. We defined multiple testing-corrected statistical significance for each trait as having a Benjamini-Hochberg false discovery rate (FDR) < 0.05. For each identified protein, an interaction term of the associated phenotype and each of the other two phenotypes would be added in the multiple linear regression models to evaluate the joint effect of phenotypes on the proteins.

To assess potential modules of proteomic markers and their phenotypic associations, we performed a principal component (PC) analysis of 73 proteins, which passed quality control procedures. We evaluated the associations for the top PCs that explained approximately 90% variance of the proteomes with age, sex, and CD4+ T-cell count separately, controlling for the same covariates described above using multiple linear regressions models.

## Results

The demographic and clinical characteristics of 87 South African PWH in ADReSS are summarized in Table 1. Our study population was 100% Black and included both men (41.4%) and women (58.6%). Participants included from the rural area were older (33.1 vs 31.1) and had higher CD4+ T-cell counts (323.5 vs 259.0 count/μL) than PWH from the urban area. All 73 inflammation-related proteins in ADReSS were correlated with at least one other protein (correlation r^2^ ≥ 0.3), with most of the correlation being positive (Figure 1). Hierarchical clustering of the 73 proteins also confirmed that a majority of proteins shared similarity with each other (Figure 1). We showed that the top 21 PCs could explain approximately 90% variance of the proteomes (Figure 2A).

**Table 1.** Demographic and clinical Characteristics of the study population.

|  | RK Khan Hospital<br>(N = 43) | Bethesda Hospital<br>(N = 44) | Overall<br>(N = 87) |
| --- | --- | --- | --- |
| Female, n (%) | 27 (62.8%) | 24 (54.5%) | 51 (58.6%) |
| Age in years,<br>mean (SD) | 31.1 (6.7) | 33.1 (9.1) | 32.1 (8.0) |
| CD4 count/μL,<br>median (IQR) | 259.0<br>(96.0-383.5) | 323.5<br>(223.5-416.5) | 290.0<br>(189.0-405.0) |

Abbreviation: Standard deviation (SD), Interquartile range (IQR)

**Figure 2.**
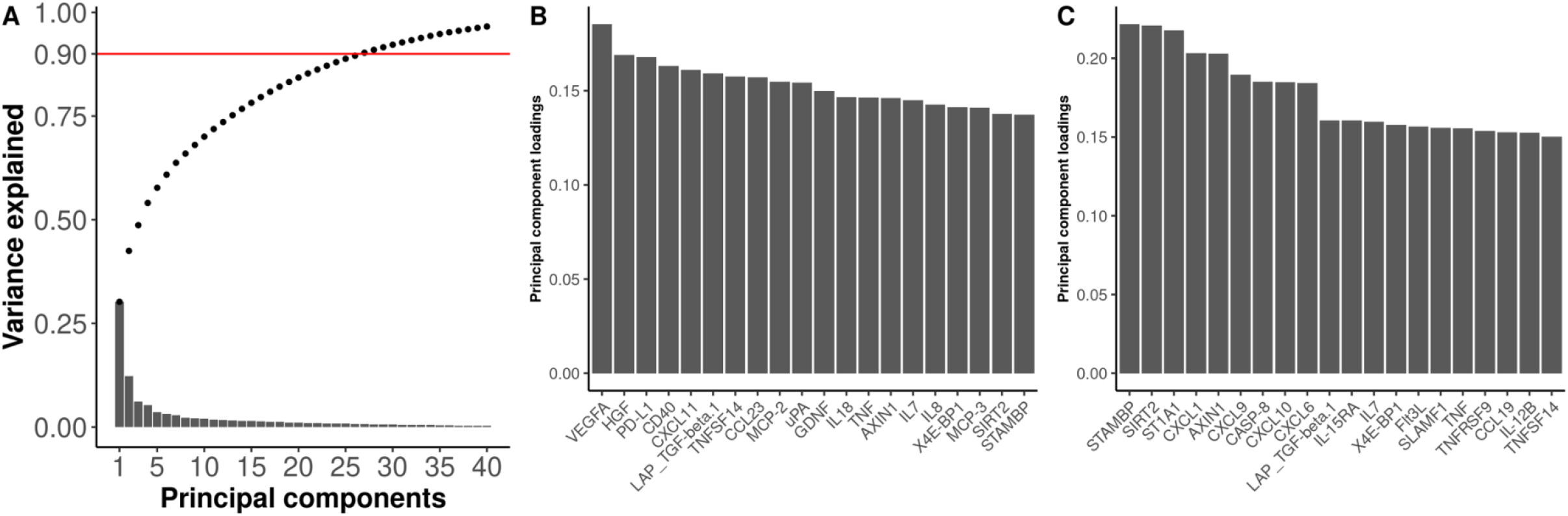
Principal component (PC) analysis of 73 proteins. **(A)** shows the variance explained by top 40 principal components. The bars indicate variance of proteomes explained by each PC and the dots show the cumulative variance explained. Red line shows 90% of variance of proteomes data could be explained by top 27 PCs cumulatively. **(B)** and **(C)** show the loadings from the 20 proteins contributed the most to PC1 and PC2 respectively.

We found 4, 18 and 19 proteins in crude associations with male sex, age and CD4 count respectively (Table 2). After adjusting for potential confounders, the number of proteins identified reduced to 1, 14 and 1, respectively (Table 3). Protein CST5 was found to be significantly associated with biological sex (male vs female) at the FDR < 0.05 level after adjusting for age and study sites, with a beta coefficient of 0.59 corresponding to a 50.5% higher NPX in the linear scale among women (95% CI: 0.29, 0.88, P-value = 2.00×10^−4^, FDR-q = 0.01). Fourteen proteins were positively associated with chronological age (Table 3) among PWH after adjusting for sex and study sites. CDCP1 showed the strongest association with chronological age, with a beta-coefficient of 0.06 corresponding to a 4.25% increase of CDCP1 abundance per year (95% CI: 0.04, 0.09, P-value = 9.65×10^−7^, FDR-q = 7.04×10^−5^). All age-associated proteins except MMP-1 were clustered together and positively correlated with each other (Figure 1 and 3). We included an additional interaction term (sex*age, age*CD4 count, sex*CD4 count) in the linear models. We found that older males had the highest NPX for CST5 compared to the other combination of sex and age (synergistic interaction, P-value = 0.04). The association between age and MCP-1 grew stronger when the CD4 count was lower (antagonistic interaction, P-value = 0.04) (Supplement table 2). Skp1-cullin-F-box (SCF)-like complex was positively associated with CD4+ T-cell counts, with an increase of 100 CD4+ T-cells/μL associated with a 14.87% increase of SCF abundance in the linear scale (beta-coefficient = 0.2, 95% CI: 0.09, 0.3, P-value = 5.49×10^−4^, FDR-q = 0.04), after controlling for age, sex and study sites. Interestingly, SCF was negatively correlated with most of the other proteins in this sample of PWH and has long clustering distance from most proteins (Figure 1).

**Table 2.** Crude phenotype-protein associations in the HIV AIDS Drug Resistance Surveillance Study with a false discovery rate-adjusted P-value less than 0.05.

| Protein | Phenotype | Beta coefficient | Fold change (%) | Standard error | P-value | FDR-q |
| --- | --- | --- | --- | --- | --- | --- |
| CST5 | Male vs Female | 0.72 | 64.57% | 0.15 | 5.58E-06 | 0.00 |
| CCL23 | Male vs Female | 0.51 | 42.37% | 0.14 | 4.86E-04 | 0.02 |
| MCP-3 | Male vs Female | 0.66 | 58.35% | 0.19 | 8.40E-04 | 0.02 |
| CDCP1 | Male vs Female | 0.69 | 61.62% | 0.21 | 1.55E-03 | 0.03 |
| CDCP1 | Age | 0.07 | 4.98% | 0.01 | 3.64E-08 | 2.66E-06 |
| SLAMF1 | Age | 0.04 | 2.97% | 0.01 | 2.09E-06 | 7.61E-05 |
| MCP-3 | Age | 0.06 | 3.89% | 0.01 | 3.52E-06 | 8.57E-05 |
| CXCL9 | Age | 0.05 | 3.42% | 0.01 | 9.90E-05 | 1.20E-03 |
| IL-15RA | Age | 0.02 | 1.34% | 0.00 | 7.09E-05 | 1.20E-03 |
| Flt3L | Age | 0.03 | 2.11% | 0.01 | 9.83E-05 | 1.20E-03 |
| CST5 | Age | 0.04 | 2.66% | 0.01 | 1.34E-04 | 1.40E-03 |
| CCL23 | Age | 0.03 | 2.21% | 0.01 | 4.66E-04 | 4.25E-03 |
| CXCL10 | Age | 0.05 | 3.20% | 0.01 | 1.33E-03 | 0.01 |
| uPA | Age | 0.02 | 1.44% | 0.01 | 1.85E-03 | 0.01 |
| MCP-2 | Age | 0.03 | 2.30% | 0.01 | 1.76E-03 | 0.01 |
| CCL25 | Age | 0.03 | 1.84% | 0.01 | 1.60E-03 | 0.01 |
| TNFRSF9 | Age | 0.03 | 1.81% | 0.01 | 2.36E-03 | 0.01 |
| MCP-1 | Age | 0.03 | 2.16% | 0.01 | 2.64E-03 | 0.01 |
| CCL11 | Age | 0.03 | 1.99% | 0.01 | 3.90E-03 | 0.02 |
| IL-10RA | Age | 0.02 | 1.51% | 0.01 | 3.97E-03 | 0.02 |
| CSF-1 | Age | 0.01 | 0.75% | 0.00 | 8.52E-03 | 0.04 |
| CCL4 | Age | 0.03 | 1.96% | 0.01 | 9.99E-03 | 0.04 |
| SCF | CD4 count | 0.21 | 15.95% | 0.05 | 7.47E-05 | 2.73E-03 |
| FGF-21 | CD4 count | -0.51 | -29.70% | 0.12 | 7.05E-05 | 2.73E-03 |
| MCP-1 | CD4 count | -0.19 | -12.46% | 0.05 | 1.57E-04 | 3.82E-03 |
| IL-15RA | CD4 count | -0.08 | -5.72% | 0.02 | 5.23E-04 | 0.01 |
| PD-L1 | CD4 count | -0.15 | -9.79% | 0.04 | 6.31E-04 | 0.01 |
| MCP-2 | CD4 count | -0.18 | -11.47% | 0.05 | 8.15E-04 | 0.01 |
| SLAMF1 | CD4 count | -0.15 | -9.73% | 0.04 | 1.45E-03 | 0.02 |
| TNF | CD4 count | -0.17 | -10.98% | 0.05 | 1.85E-03 | 0.02 |
| CCL23 | CD4 count | -0.14 | -9.07% | 0.04 | 2.66E-03 | 0.02 |
| IL18 | CD4 count | -0.18 | -11.69% | 0.06 | 3.08E-03 | 0.02 |
| CSF-1 | CD4 count | -0.06 | -4.05% | 0.02 | 3.51E-03 | 0.02 |
| CXCL10 | CD4 count | -0.21 | -13.26% | 0.07 | 4.20E-03 | 0.03 |
| CCL19 | CD4 count | -0.17 | -10.97% | 0.06 | 0.01 | 0.04 |
| IL6 | CD4 count | -0.19 | -12.63% | 0.07 | 0.01 | 0.04 |
| MMP-1 | CD4 count | -0.22 | -13.94% | 0.08 | 0.01 | 0.04 |
| CCL3 | CD4 count | -0.29 | -18.48% | 0.11 | 0.01 | 0.04 |
| IFN-gamma | CD4 count | -0.27 | -17.15% | 0.10 | 0.01 | 0.04 |
| IL-17C | CD4 count | -0.14 | -9.21% | 0.05 | 0.01 | 0.04 |
| CXCL9 | CD4 count | -0.16 | -10.49% | 0.06 | 0.01 | 0.0497 |

**Table 3.** Adjusted phenotype-protein associations in the HIV AIDS Drug Resistance Surveillance Study with a false discovery rate-adjusted P-value less than 0.05.

| Protein | Phenotype | Beta coefficient | Fold change (%) | Standard error | P-value | FDR-q |
| --- | --- | --- | --- | --- | --- | --- |
| CST5 | Male vs Female | 0.59 | 50.52 | 0.15 | $2.00 \times 10^{-4}$ | 0.01 |
| | Age | 0.03 | 2.10 | 0.01 | $4.57 \times 10^{-3}$ | 0.04 |
| SLAMF1 | Age | 0.04 | 2.81 | 0.01 | $1.05 \times 10^{-5}$ | $3.84 \times 10^{-4}$ |
| MCP-3 | Age | 0.05 | 3.53 | 0.01 | $1.35 \times 10^{-4}$ | $2.48 \times 10^{-3}$ |
| IL-15RA | Age | 0.02 | 1.40 | 0.01 | $1.66 \times 10^{-4}$ | $2.48 \times 10^{-3}$ |
| Flt3L | Age | 0.03 | 2.10 | 0.01 | $1.70 \times 10^{-4}$ | $2.48 \times 10^{-3}$ |
| CXCL9 | Age | 0.04 | 2.81 | 0.01 | $1.33 \times 10^{-3}$ | 0.02 |
| MCP-1 | Age | 0.03 | 2.10 | 0.01 | $4.06 \times 10^{-3}$ | 0.04 |
| CDCP1 | Age | 0.06 | 4.25 | 0.01 | $9.65 \times 10^{-7}$ | $7.04 \times 10^{-5}$ |
| CXCL10 | Age | 0.04 | 2.81 | 0.01 | $4.76 \times 10^{-3}$ | 0.04 |
| uPA | Age | 0.02 | 1.40 | 0.01 | $6.59 \times 10^{-3}$ | 0.04 |
| TNFRSF9 | Age | 0.02 | 1.40 | 0.01 | $6.09 \times 10^{-3}$ | 0.04 |
| MMP-1 | Age | 0.04 | 2.81 | 0.02 | $8.60 \times 10^{-3}$ | 0.05 |
| CCL23 | Age | 0.02 | 1.40 | 0.01 | $9.53 \times 10^{-3}$ | 0.05 |
| MCP-2 | Age | 0.03 | 2.10 | 0.01 | $9.29 \times 10^{-3}$ | 0.05 |
| SCF | CD4 | 0.2 | 14.87 | 0.05 | $5.49 \times 10^{-4}$ | 0.04 |

**Figure 3.**
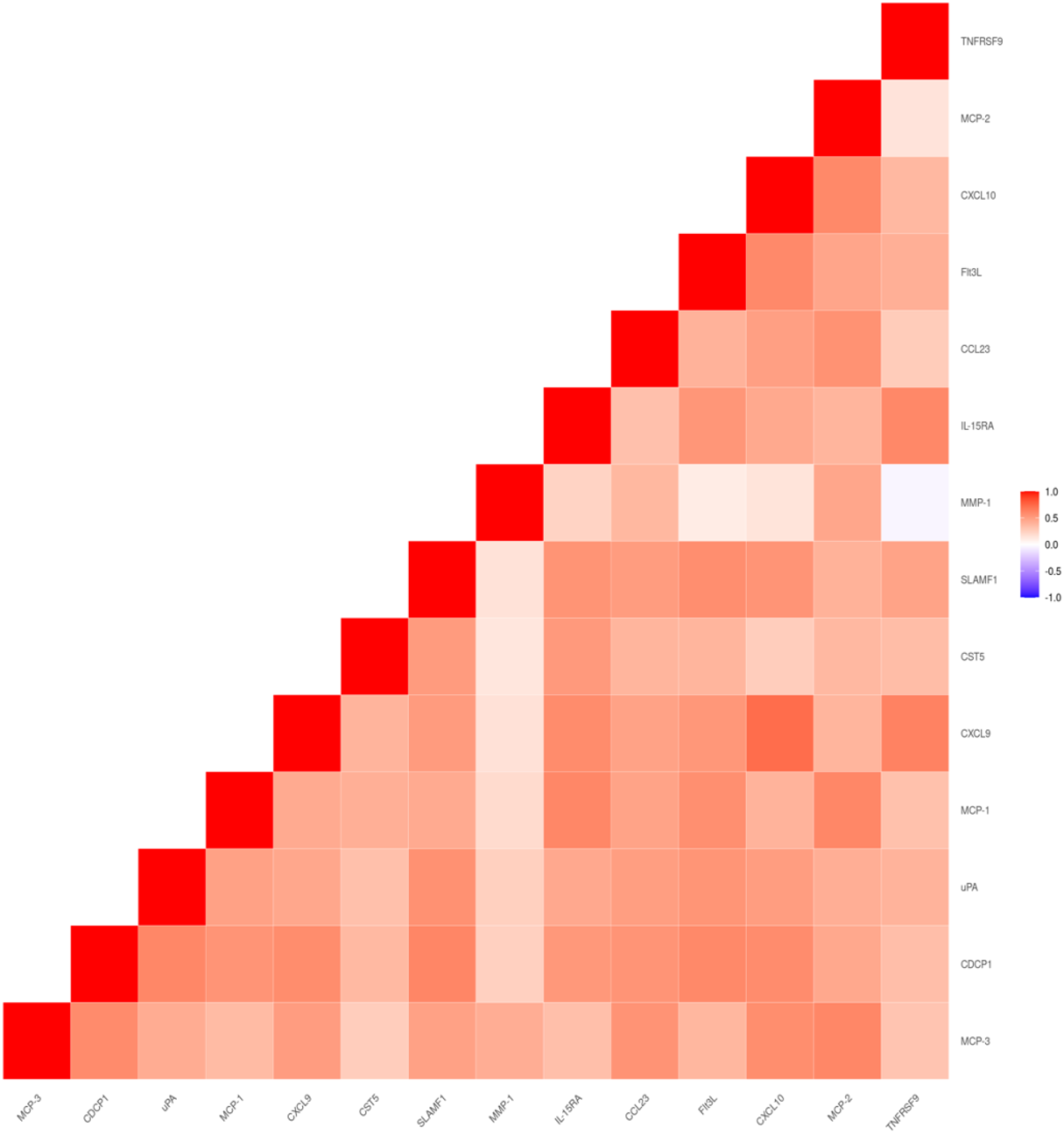
Heatmap showing Spearman correlation between 14 age-associated proteins.

After adjusting for sex, we found that PC1 and PC2 calculated from the proteomes were associated with age, having nominal P-values of 8.41×10^−3^ and 3.09×10^−3^ (Table 4). Additionally, CD4+ T-cell count was also associated with PC2 (P-value = 0.02). However, these associations were not statistically significant after adjusting for multiple testing. The loadings of proteins for each PC were the coefficients of the linear combination of the proteins from which the PC was constructed (Figure 2 B & C and Supplement table 2), and a positive loading indicates that a protein contributes to some degree to the PCs. We noted that age-associated proteins including CXCL9, CXCL10, SLAMF1 and TNFSF9 contributed to PC2 (Figure 2C) while CD4+ cell count-associated protein SCF has a loading of 0.06 for PC2 (Supplement table 2).

**Table 4.**
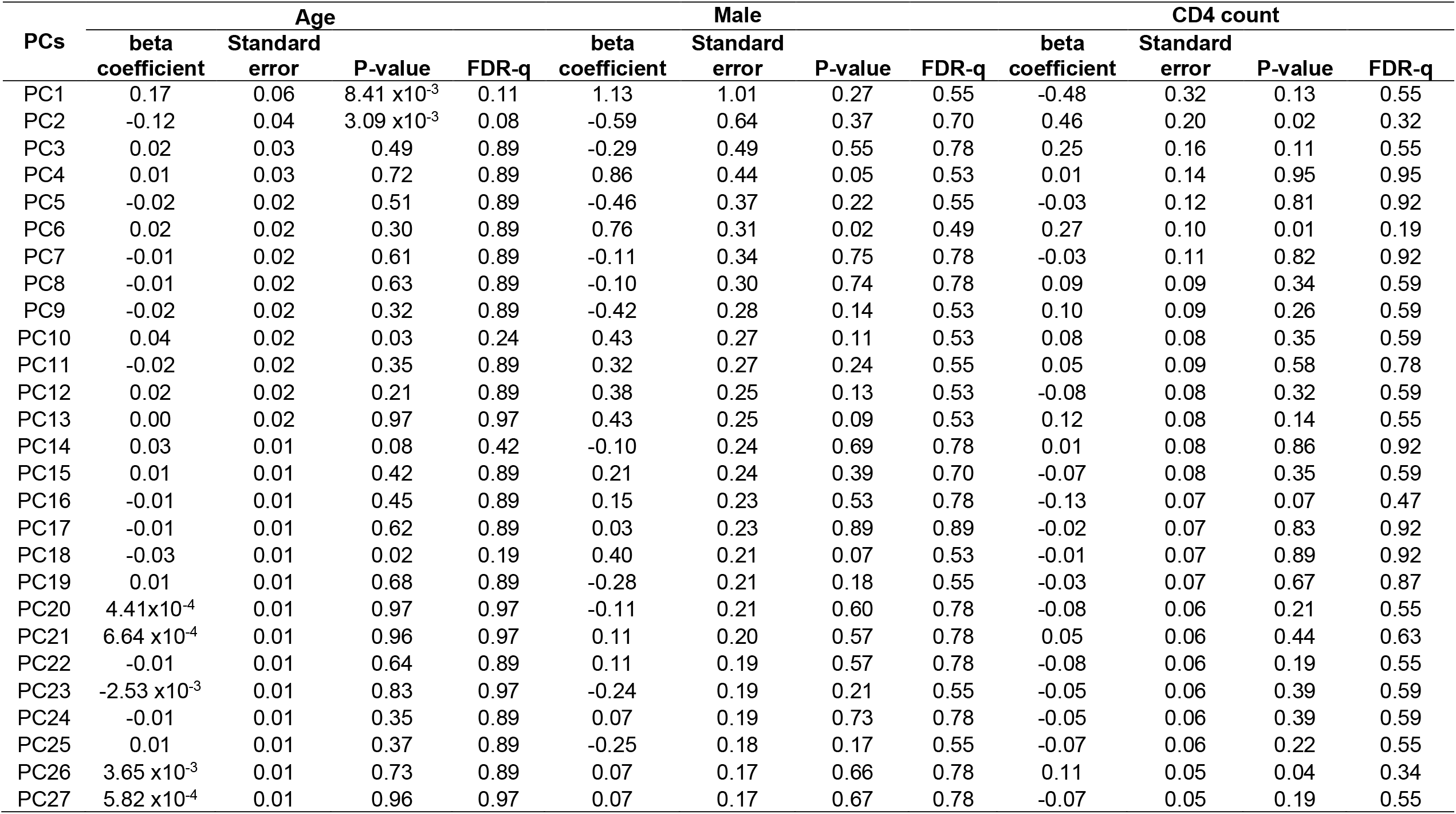
**Associations of chronological age, sex (female vs male) and CD4 count with top 27 principal components (PC) which explained total 90% of the variation of inflammation related proteomes. Beta coefficients for CD4 count indicates changes of PCs associated with 100-unit increase of CD4 count.**

| PCs | Age |  |  |  | Male |  |  |  | CD4 count |  |  |  |
| --- | --- | --- | --- | --- | --- | --- | --- | --- | --- | --- | --- | --- |
|  | beta<br>coefficient | Standard<br>error | P-value | FDR-q | beta<br>coefficient | Standard<br>error | P-value | FDR-q | beta<br>coefficient | Standard<br>error | P-value | FDR-q |
| PC1 | 0.17 | 0.06 | 8.41 x10<sup>-3</sup> | 0.11 | 1.13 | 1.01 | 0.27 | 0.55 | -0.48 | 0.32 | 0.13 | 0.55 |
| PC2 | -0.12 | 0.04 | 3.09 x10<sup>-3</sup> | 0.08 | -0.59 | 0.64 | 0.37 | 0.70 | 0.46 | 0.20 | 0.02 | 0.32 |
| PC3 | 0.02 | 0.03 | 0.49 | 0.89 | -0.29 | 0.49 | 0.55 | 0.78 | 0.25 | 0.16 | 0.11 | 0.55 |
| PC4 | 0.01 | 0.03 | 0.72 | 0.89 | 0.86 | 0.44 | 0.05 | 0.53 | 0.01 | 0.14 | 0.95 | 0.95 |
| PC5 | -0.02 | 0.02 | 0.51 | 0.89 | -0.46 | 0.37 | 0.22 | 0.55 | -0.03 | 0.12 | 0.81 | 0.92 |
| PC6 | 0.02 | 0.02 | 0.30 | 0.89 | 0.76 | 0.31 | 0.02 | 0.49 | 0.27 | 0.10 | 0.01 | 0.19 |
| PC7 | -0.01 | 0.02 | 0.61 | 0.89 | -0.11 | 0.34 | 0.75 | 0.78 | -0.03 | 0.11 | 0.82 | 0.92 |
| PC8 | -0.01 | 0.02 | 0.63 | 0.89 | -0.10 | 0.30 | 0.74 | 0.78 | 0.09 | 0.09 | 0.34 | 0.59 |
| PC9 | -0.02 | 0.02 | 0.32 | 0.89 | -0.42 | 0.28 | 0.14 | 0.53 | 0.10 | 0.09 | 0.26 | 0.59 |
| PC10 | 0.04 | 0.02 | 0.03 | 0.24 | 0.43 | 0.27 | 0.11 | 0.53 | 0.08 | 0.08 | 0.35 | 0.59 |
| PC11 | -0.02 | 0.02 | 0.35 | 0.89 | 0.32 | 0.27 | 0.24 | 0.55 | 0.05 | 0.09 | 0.58 | 0.78 |
| PC12 | 0.02 | 0.02 | 0.21 | 0.89 | 0.38 | 0.25 | 0.13 | 0.53 | -0.08 | 0.08 | 0.32 | 0.59 |
| PC13 | 0.00 | 0.02 | 0.97 | 0.97 | 0.43 | 0.25 | 0.09 | 0.53 | 0.12 | 0.08 | 0.14 | 0.55 |
| PC14 | 0.03 | 0.01 | 0.08 | 0.42 | -0.10 | 0.24 | 0.69 | 0.78 | 0.01 | 0.08 | 0.86 | 0.92 |
| PC15 | 0.01 | 0.01 | 0.42 | 0.89 | 0.21 | 0.24 | 0.39 | 0.70 | -0.07 | 0.08 | 0.35 | 0.59 |
| PC16 | -0.01 | 0.01 | 0.45 | 0.89 | 0.15 | 0.23 | 0.53 | 0.78 | -0.13 | 0.07 | 0.07 | 0.47 |
| PC17 | -0.01 | 0.01 | 0.62 | 0.89 | 0.03 | 0.23 | 0.89 | 0.89 | -0.02 | 0.07 | 0.83 | 0.92 |
| PC18 | -0.03 | 0.01 | 0.02 | 0.19 | 0.40 | 0.21 | 0.07 | 0.53 | -0.01 | 0.07 | 0.89 | 0.92 |
| PC19 | 0.01 | 0.01 | 0.68 | 0.89 | -0.28 | 0.21 | 0.18 | 0.55 | -0.03 | 0.07 | 0.67 | 0.87 |
| PC20 | 4.41x10<sup>-4</sup> | 0.01 | 0.97 | 0.97 | -0.11 | 0.21 | 0.60 | 0.78 | -0.08 | 0.06 | 0.21 | 0.55 |
| PC21 | 6.64 x10<sup>-4</sup> | 0.01 | 0.96 | 0.97 | 0.11 | 0.20 | 0.57 | 0.78 | 0.05 | 0.06 | 0.44 | 0.63 |
| PC22 | -0.01 | 0.01 | 0.64 | 0.89 | 0.11 | 0.19 | 0.57 | 0.78 | -0.08 | 0.06 | 0.19 | 0.55 |
| PC23 | -2.53 x10<sup>-3</sup> | 0.01 | 0.83 | 0.97 | -0.24 | 0.19 | 0.21 | 0.55 | -0.05 | 0.06 | 0.39 | 0.59 |
| PC24 | -0.01 | 0.01 | 0.35 | 0.89 | 0.07 | 0.19 | 0.73 | 0.78 | -0.05 | 0.06 | 0.39 | 0.59 |
| PC25 | 0.01 | 0.01 | 0.37 | 0.89 | -0.25 | 0.18 | 0.17 | 0.55 | -0.07 | 0.06 | 0.22 | 0.55 |
| PC26 | 3.65 x10<sup>-3</sup> | 0.01 | 0.73 | 0.89 | 0.07 | 0.17 | 0.66 | 0.78 | 0.11 | 0.05 | 0.04 | 0.34 |
| PC27 | 5.82 x10<sup>-4</sup> | 0.01 | 0.96 | 0.97 | 0.07 | 0.17 | 0.67 | 0.78 | -0.07 | 0.05 | 0.19 | 0.55 |

## Discussion

Our study investigated proteomic associations with demographic and clinical characteristics of black PWH prior to initiating ART in South Africa, a population with a high burden of age-related comorbidities. The elevated abundance of 14 soluble inflammatory protein markers were associated with older age. Among these age-associated proteins, MCP-1, CXCL9, CCXCL19 and CDCP1 were previously found to be associated with age among 192 virally suppressed PWH^16^. Furthermore, all but two proteins (CCL23, uPA) showed significantly increased expression for older individuals among 416 healthy participants^16^, and 5 of them including SLAMF1, IL-15RA, FLT3L, CXCL9 and CXCL10, have been previously reported to be associated with older age in a large-scale proteomes analysis of 2,925 plasma proteins and 4,263 participants without HIV^19^. This suggests there may be a shared proteomic pattern between PWH and people without HIV when they are growing older. Many current research efforts have focused on developing age-associated biomarkers, such as epigenetic “clocks” based on changes in DNA methylation level^7^. Incorporating information from these age-associated proteins into the development of such “clocks” may improve early prediction of unhealthy aging. We also found a synergistic interaction between male sex and older age accounting for excessive expression of CST5, and an interaction between low CD4 count and older age accounting for excessive expression of MCP-1.

Additionally, age-associated proteins CST5, CCL23, SLAMF1, MMP-1, MCP-1, and CDCP1 were shown to be elevated among PWH compared to people living without HIV regardless of long-term use of ART^6,12^. This suggests HIV infection may lead to unhealthy aging through similar pathways as advanced chronological age or accelerated biological aging, which highlights the need for clinical interventions or anti-inflammatory drugs targeting those PWH who are aging. CCL23, SLAMF1 and MCP-1 are involved in monocyte activation, which could lead to sustained inflammation among PWH^6,12^ and were suggested to be involved in the pathogensis of several age-related diseases, such as cardiovascular diseases (CVD)^20^, chronic kidney disease^21^, rheumatoid arthritis and atopic dermatitis^6,22,23^. CCL23 was shown to be positively associated with the presence of coronary plaque among PWH^24^, and genetic variants in the *SLAMF1* gene that regulate the abundance of SLAMF1 protein were associated with memory decline^25^. MMP-1 functions primarily to degrade several types of collagens and plays an important role in skin aging^26^. Elevated MMP-1 was also associated diverse cancer types^27,28^, providing further evidence of its role in diseases related to aging. Further, increased risks of HIV comorbidities such as hypertension, diabetes, and CVD and cancer were observed among older PWH^4,8^. Previous research has also shown that male and female PWH might bear different but equally heavy burdens of HIV comorbidities^29^. For example, recent data showed that CVD mortality was higher among women with HIV (rate ratio = 2.24) compared to men with HIV (rate ratio = 1.23)^30^, but men with HIV had a higher prevalence of hypertension than women with HIV^31^. These observations might link sexual-dimorphic protein levels with differential chronic disease outcomes among PWH. Further studies are warranted to investigate the relationship between these sex-/age-associated proteins and HIV comorbidities.

Our study showed a positive association between CD4+ T-cell counts and SCF complex, which is a multi-protein E3 ubiquitin ligase complex and has a vital role in the ubiquitination of proteins involved in the cell cycle^32^. Ubiquitin machinery involving the SCF complex is an essential pathway through which cyclin F could possibly regulate HIV-1 Viral Infectivity Factor^33^, which might lead to the increase of CD4+ T-cell counts among PWH. However, whether SCF complex plays a role in the development of comorbidities among PWH remained unknown.

The PC2 of proteomic data is associated with CD4+ T-cell counts (P-value of 0.02), although not statistically significant after multiplicity testing. The loading of only CD4+ T-cell counts-associated protein SCF for PC2 was only 0.06, suggesting that the CD4+ T-cell count-PC2 association might have arisen from individual or combined signals of multiple other proteins, including some of the age-associated proteins. This finding showed the potential benefits of using dimension reduction such as PC analysis to account for the relatedness between proteins, reduce the multiple testing burden, and increase the proteomic study power.

In this exploratory study, we identified proteomic associations with demographic and clinical characteristics of South African PWH prior to initiating ART. Since genetic and environmental factors enormously impact on the proteomic profiling^34^, the proteomic patterns and consequences of aging might differ across populations^35^. Additionally, studies conducted in European populations might not apply to individuals of other ancestries or from low/middle-income countries. Our study demonstrated the value of such proteomic studies among underrepresented populations to fill these knowledge gaps. We also acknowledge a few limitations in the present study. First, the relatively small sample size limited our study power. Future replication studies in larger samples are needed to validate these findings. Additionally, the cross-sectional study design restricted the capacity to determine causal inferences on the relationships between phenotypes.

In conclusion, this study highlighted multiple proteins associated with demographic and clinical characteristics of ART-naïve PWH living in South Africa, demonstrating the promise of using proteomic data to evaluate and study the inflammatory status among PWH. Several inflammatory proteins have shown a strong association with age and could be the next targets to investigate age-related comorbidities and predict patterns of aging for PWH.

## Data Availability

The data presented in this study are available on request from the corresponding author.

## Data Availability Statements

The data presented in this study are available on request from the corresponding author.

## Acknowledgement

This study is funded by NIDDK (R01 DK125187). VCM received support from the Emory CFAR (P30 AI050409) and NIH/NIAID (R01 AI098558).

